# Informant-rated change in personality traits, psychological distress, well-being, and social connection with dementia

**DOI:** 10.1101/2023.08.18.23294273

**Authors:** Angelina R. Sutin, Martina Luchetti, Yannick Stephan, Antonio Terracciano

## Abstract

**Objectives:** Studies of retrospective personality change with dementia consistently find caregivers report large changes in personality (e.g., increases in neuroticism) of their care recipients compared to before dementia. This work seeks to replicate the established pattern of personality change, extend it to change in psychological distress, well-being, and social connection, and evaluate whether changes vary by stage of dementia. **Methods:** Caregivers of people with dementia (*N*=194) reported on the psychological and social health of their care recipient currently and how they were before they developed dementia. Personality was measured as five factor model traits. Psychological distress was measured as symptoms of depression and anxiety, perceived stress, and pessimism. Psychological well-being was measured as purpose in life, life satisfaction, happiness, self-efficacy, and optimism. Social connection was measured as loneliness, belonging, social support, and social strain. **Results:** There were substantial increases in neuroticism (*d*=1.10) and decreases in the other four personality traits (*d* range=-.82 to −1.31). There were significant increases in psychological distress (e.g., *d*=1.05 for depression) and substantial decreases in well-being (e.g., *d*=-1.07 for purpose in life) and social connection (e.g., *d*=-1.09 for belonging). Change was apparent across dementia stage and generally larger in more severe dementia. **Discussion:** In addition to personality, there are large retrospective changes in psychological distress, well-being, and social connection with dementia. These quantitative findings complement clinical observations of the natural history of psychosocial changes in people with dementia, and can inform families, clinicians, and researchers on commonly observed changes and improve interventions to mitigate dementia burden.

---

Significant changes in psychological function (e.g., mood, personality) have been a hallmark of cognitive impairment since at least the time of Alzheimer’s first patient, Auguste D.^1^ Today, such changes are a core clinical criterion for a diagnosis of Alzheimer’s disease or related dementia (ADRD).^2^ These changes likely result in part from the widespread neurological changes and neurodegeneration caused by dementia.^3^ Changes in lifestyle and social interactions that often accompany cognitive decline and dementia may also contribute to these changes.^4^ Psychological changes persist throughout the course of the disease and are one of the most difficult aspects of caregiving, especially when problematic behaviors are manifested as behavioral and psychological symptoms of dementia (BPSD).^5^ Much of the focus on psychological function in dementia has been on BPSD, which are defined in part by their new emergence during the disease,^6^ rather than on how aspects of psychological function may change compared to before the onset of dementia.

There is, however, a robust literature on change in five factor model personality traits (FFM) with dementia.^7–9^ One common approach is to ask caregivers of the individual with dementia to rate the personality of their care-recipient both currently and prior to dementia.^10^ Such retrospective ratings have yielded very consistent perceived changes in personality: A meta-analysis^11^ of 18 samples found that informants perceived the individual with dementia to increase substantially in neuroticism (the tendency to experience negative emotions and vulnerability to stress) and decrease substantially in extraversion (the tendency to be sociable and outgoing) and conscientiousness (the tendency to be organized, disciplined, and responsible) and also to have more modest declines in openness (the tendency to be creative and unconventional) and agreeableness (the tendency to be straightforward and trusting). There are, however, numerous other aspects of psychological and social function that are likely susceptible to change during dementia that have yet to be examined with this method. The present research thus uses this approach established to examine change in personality and applies it to other aspects of psychological function and social connection. Specifically, in addition to the five major personality traits, this study includes multiple aspects of psychological distress (anxiety, depression, stress, pessimism), psychological well-being (purpose in life, life satisfaction, happiness, self-efficacy, optimism), and social connection (loneliness, belonging, social support, social strain).

Change in personality, psychological distress and well-being, and social connection may vary by stage of dementia. Previous research on retrospective personality change has generally not had sample sizes large enough to evaluate change by dementia stage (average *N*=28^11^). There is, however, evidence that retrospective personality change in mild cognitive impairment (MCI) is smaller in magnitude than change during dementia.^11^ Further, longitudinal work on BPSD has found that symptoms tend to get worse over time^12^ and across dementia stage.^13^ This pattern suggests that there is likely to be more substantial change in psychological expression as neurodegeneration progresses across the continuum of the disease. This progression may result in greater changes in psychological function and social connection as the disease advances.

The purpose of this research is to replicate past findings on retrospective change in personality and to use this methodology to extend understanding of changes with dementia to other aspects of psychological function and social connection. Given the previous meta-analysis found very consistent retrospective changes in personality reported in the published literature,^11^ we expect the same pattern of change (increases in neuroticism and its facets and declines in the other domains and their facets). We also expect to see a similar pattern of change for the other dimensions such that psychological distress (e.g., depression) and negative aspects of social connection (e.g., loneliness) will increase and positive aspects of psychological well-being (e.g., purpose in life) and social connection (e.g., social support) will decrease. With the largest sample to date with primary data, we also examine whether retrospective change varies by stage of dementia. We expect changes to be more pronounced in moderate and severe dementia than mild dementia because previous work has found smaller retrospective changes in individuals with mild cognitive impairment compared to dementia.^11^

## Method

### Participants and Procedure

Participants were caregivers of adults with dementia. Participants were recruited through Dynata’s proprietary online panel of caregivers and tested in February 2023. Inclusion criteria were currently caring for an adult 60 years or older with dementia, knew the care recipient before the diagnosis of dementia, and living in the United States. Of the caregivers who met these criteria who completed the survey (*n*=201), those who took less than five minutes to do so were excluded from the analysis (*n*=7). A total of 194 caregivers of individuals with dementia were included in the analytic sample.

The protocol was approved by the institutional review board at the Florida State University (#STUDY00001134) and carried out in accordance with the Declaration of Helsinki. All participants provided informed consent before completing the survey.

### Measures

For each scale below, participants rated the items twice: Once as they perceived the person with dementia prior to diagnosis and once as they perceived the person now. The order of presentation (retrospective versus current) was randomized across participants. Below, we give example items for each scale in the present tense that was used to measure the care recipient’s current psychological function and social connection. Items were worded in the past tense for retrospective ratings prior to diagnosis (see Supplemental Material for an example of the survey). Items were also worded based on the caregiver’s preferred pronoun to refer to their care-recipient (he/she/they). Unless otherwise noted, items were rated on a scale from 1 (*strongly disagree*) to 5 (*strongly agree*).

#### Personality traits

Five-factor personality traits were measured with the 30-item version of the Big Five Inventory-2 (BFI-2-S).^14^ Participants rated items that completed the sentence stem, “He/She/They is someone who…” The items measured neuroticism (e.g., worries a lot), extraversion (e.g., is outgoing, sociable), openness (e.g., is complex, a deep thinker), agreeableness (e.g., is compassionate, has a soft heart), and conscientiousness (e.g., is systematic, likes keeping things in order). Items were reverse scored in the direction of the trait label when necessary and the mean taken across items for each domain. In addition to the five broad domains, three facets for each trait were also scored: anxiety, depression, and emotional volatility (neuroticism), sociability, assertiveness, and energy level (extraversion), intellectual curiosity, aesthetic sensitivity, and creative imagination (openness), compassion, respectfulness, and trust (agreeableness), and organization, productiveness, and responsibility (conscientiousness).

#### Psychological distress

The Patient Health Question-4 (PHQ-4)^15^ was used to measure anxiety with two items (e.g., “Feeling nervous, anxious or on edge”) and depression with two items (e.g., “Feeling down, depressed or hopeless”) on a scale from 0 (*not at all*) to 3 (*nearly all the time*). The sum of the two items was taken separately for anxiety and depression. Perceived stress was measured with the item, “How would he/she/they rate the amount of stress in his/her/their life?” on a scale from 1 (*no stress*) to 6 (*extreme stress*).^16^

#### Psychological well-being

Purpose in life was measured with the 4-item (e.g., “life has purpose”) NIH PROMIS measure of purpose in life.^17^ Items were reverse scored when necessary and the mean taken in the direction of higher purpose. Life satisfaction was measured with the item, “He/She/They is someone who is satisfied with his/her/their life.” Happiness was measured with the item, “He/She/They is someone who feels happy in general.” Self-efficacy was measured with the item, “He/She/They is someone who is confident in his/her/their his/her/their ability to solve problems that he/she/they may face in life.” The 6-item Life Orientation Test-Revised (LOTR)^18^ scale measured optimism with three items (e.g., “is always optimistic about his/her/their future”) and pessimism with three items (e.g., “feels if something could go wrong for him/her/them it will”). The mean of the items was taken separately for optimism and pessimism in the direction of the labels.

#### Social connection

Social support was measured with four items (e.g., “He/She/They is someone who has someone to talk with when he/her/they need to talk”) and the mean taken in the direction of greater social support.^19^ Social strain was measured with the item, “He/She/They is someone who has people in his/her/their life who criticize him/her/them.”^19^ Belonging was measured with the item, “How would he/she/they describe his/her/their sense of belonging to his/her/their local community?” on a scale from 1 (*very weak*) to 4 (*very strong*). Loneliness was measured with the 3-item short version of the UCLA Loneliness scale.^20^ Items (e.g., “How much of the time does he/she/they feel he/she/they lack companionship?”) were rated from 1 (*hardly ever or never*) to 3 (*often*) and the mean taken in the direction of greater loneliness.

#### Stage of dementia

Caregivers specified the stage of dementia of their care recipient. The stages were defined as mild dementia (memory loss of recent events, getting lost or misplacing objects, but symptoms are not always easy to notice, and many people with mild dementia remain relatively independent), moderate dementia (increasing confusion, increased memory loss, symptoms are noticeable, and help is often needed with routine tasks), or severe dementia (loss of physical capability or to communicate, a need for full-time daily assistance with tasks, such as eating and dressing).

### Statistical Approach

Consistent with previous work on retrospective change in personality with dementia,^11^ change in all aspects of psychological and social function was evaluated with a paired-sample t-test and quantified with Cohen’s *d*. To examine whether change varied by stage of dementia, we first calculated a change score for each dimension (current – before dementia).^21^ A one-way ANOVA was then run for each dimension to test whether the change score varied by stage of dementia (mild, moderate, severe). If the ANOVA was significant, a Tukey post-hoc analysis was used to identify which stages were significantly different from each other.

## Results

The 194 individuals with dementia reported on by their caregivers in this study had a mean age of 81.79 (SD=9.06), were 65% (*n*=126) female, 90% (*n*=176) white, and 27% (*n*=53) had at least a college degree (Table 1). Caregivers had a mean age of 60.37 (SD=12.99), were 62% (*n*=120) female, 93% (*n*=180) white, and were the spouse (22%; *n*=42), adult child (47%; *n*=91), other family member (22%; *n*=43) or had another relationship with the care recipient (9%, *n*=18). Caregivers reported that their care recipient was in the mild (24%; *n*=46), moderate (64%; *n*=124), or severe (12%; *n*=24) stage of dementia.

**Table 1.** Sociodemographic Characteristics of Participants with Dementia and their Caregivers.

|  | Mean (standard deviation)<br>or Percentage (n) |
| --- | --- |
| Care-recipient with dementia |  |
| Age in years | 81.79 (9.06) |
| Sex (female) | 65% (126) |
| Race (white) | 91% (176) |
| Education (college degree or higher) | 27% (53) |
| Stage of dementia |  |
| Mild | 24% (46) |
| Moderate | 64% (124) |
| Severe | 12% (24) |
| Caregiver |  |
| Age in years | 60.37 (13.99) |
| Sex (female) | 62% (120) |
| Race (white) | 93% (180) |
| Education (college degree or higher) | 52% (101) |
| Relationship |  |
| Spouse | 22% (42) |
| Adult child | 47% (91) |
| Other family | 22% (43) |
| Other relationship | 9% (18) |

*Note.* N=194.

Means for personality traits before and during dementia are in Table 2. Consistent with the literature on retrospective change in personality,^11^ the largest changes observed were the substantial increase in neuroticism and the substantial decrease in both extraversion and conscientiousness. Also consistent with this literature, openness and agreeableness decreased. To quantify the similarity of findings between the current study and the published meta-analytic findings, we computed a (double-entry) correlation across the five traits and found *r*=.89, *p*<.001, which indicated a replicable pattern of associations. Changes in the facets were similar to changes observed at the domain level (range = −.44 [aesthetic sensitivity, facet of openness] to −1.27 [responsibility, facet of conscientiousness]).

**Table 2.**
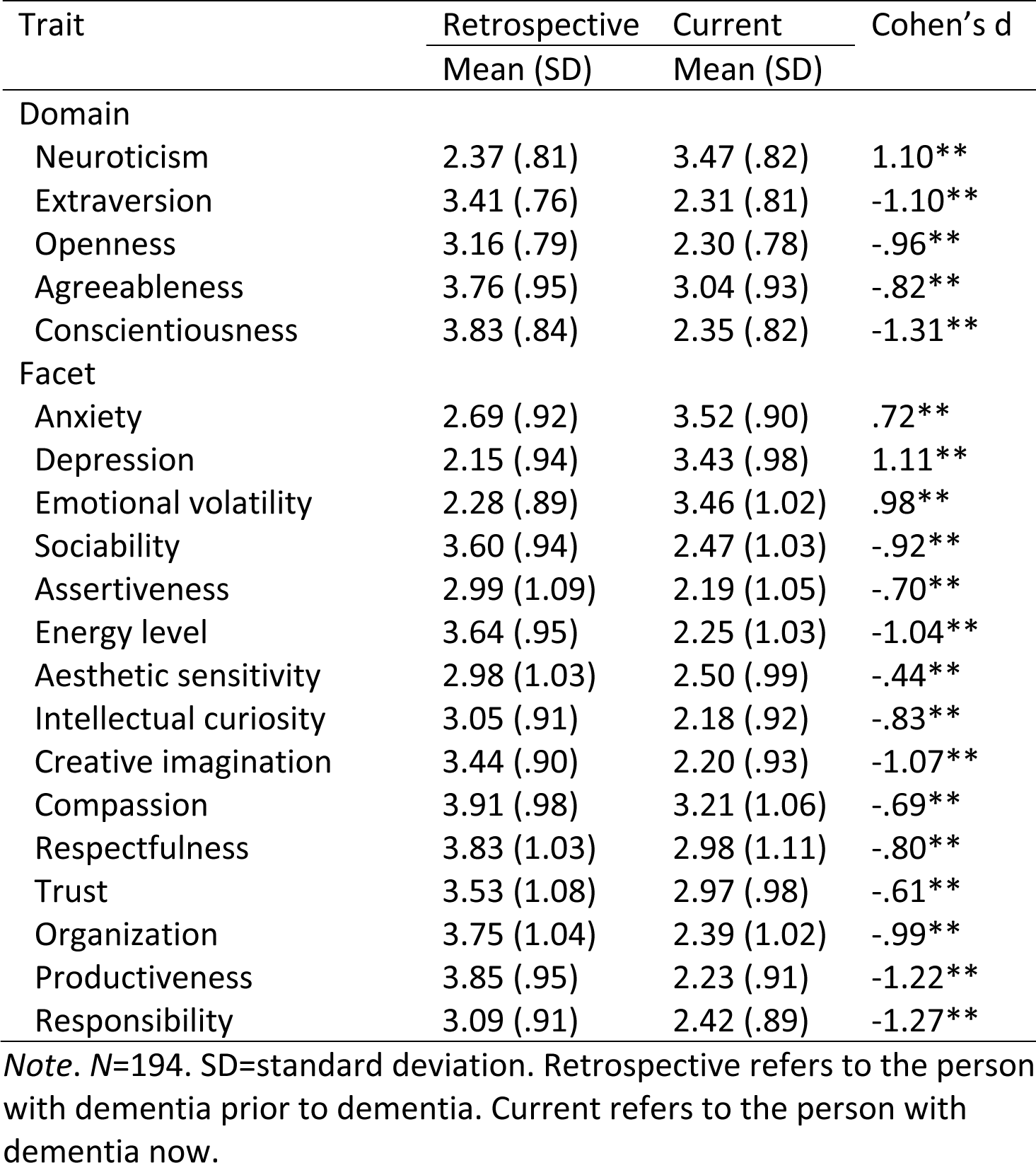
Retrospective Change in Personality Domains and Facets.

There was evidence of retrospective change in all measures of psychological distress, psychological well-being, and social connection (Table 3). Caregivers perceived that their care recipient had substantial increases in all aspects of psychological distress assessed in this study: anxiety, depression, stress, and pessimism. They also perceived significant declines in purpose in life, life satisfaction, happiness, self-efficacy, and optimism. These changes extended to aspects of social connection, although with more variability in size of the perceived change. That is, the retrospective changes ranged from .09 for social strain to −1.09 for belonging. Across all aspects of psychological function and social connection, the largest change was the decrease in self-efficacy and belonging (both *ds*=-1.09); depressive symptoms and purpose in life also had a *d* greater than 1.0. In contrast, the smallest change was the increase in social strain (*d*=.09).

**Table 3.** Retrospective change in psychological distress, well-being and social connection.

|  | Retrospective<br>Mean (SD) | Current<br>Mean (SD) | Cohen's *d* |
| --- | --- | --- | --- |
| Psychological distress |  |  |  |
| PHQ depression | 1.26 (1.61) | 3.13 (1.76) | 1.05** |
| PHQ anxiety | 1.53 (1.57) | 2.73 (1.73) | .71** |
| Perceived stress | 2.59 (1.02) | 3.22 (1.31) | .47** |
| Pessimism | 2.36 (1.03) | 3.08 (1.03) | .68** |
| Psychological well-being |  |  |  |
| Purpose in life | 3.88 (.93) | 2.51 (1.11) | -1.07** |
| Life satisfaction | 3.97 (1.08) | 2.55 (1.22) | -.96** |
| Happiness | 3.97 (1.06) | 2.76 (1.22) | -.91** |
| Self-efficacy | 3.84 (1.04) | 2.18 (1.22) | -1.09** |
| Optimism | 3.72 (.90) | 2.68 (1.03) | -.88** |
| Social connection |  |  |  |
| Loneliness | 1.48 (.60) | 1.88 (.68) | .62** |
| Belonging | 2.78 (.87) | 1.69 (.75) | -1.09** |
| Social support | 4.17 (.82) | 3.95 (.93) | -.26** |
| Social strain | 2.41 (1.24) | 2.52 (1.30) | .09** |

*Note.* *N*=194. SD=standard deviation. Retrospective refers to the person with dementia prior to dementia. Current refers to the person with dementia now.

Retrospective change scores for each dimension by stage of dementia is in Supplemental Table S1. Change by dementia stage quantified with Cohen’s *d* is in Figure 1. Change was apparent in all dimensions in each stage of dementia. There were a few patterns worth noting. First, in absolute terms, there was a general linear trajectory of mean change such that retrospective changes were larger at each stage of dementia: mean absolute *d*=.76 for mild dementia, mean absolute *d*=.88 for moderate dementia, and absolute *d*=1.14 for severe dementia. Second, changes were generally largest in more severe dementia cases, with some changes twice as large in severe cases compared to mild cases (e.g., stress, optimism). Third, when there were differences in change across stage of dementia, there was generally (but not always) more change between the moderate and severe stages than between the mild and moderate stages of dementia. Fourth, there were only a few cases in which there was significant difference in change across all three stages. Specifically, conscientiousness, particularly its productiveness facet, and purpose in life had significantly different decreases that got progressively and significantly larger at each stage of dementia. Finally, social strain had a different pattern than the other psychological and social factors: Caregivers perceived significant increases in the mild and moderate stages of dementia but a significant decrease in the severe stage relative to before the care recipient developed dementia.

**Figure 1.**
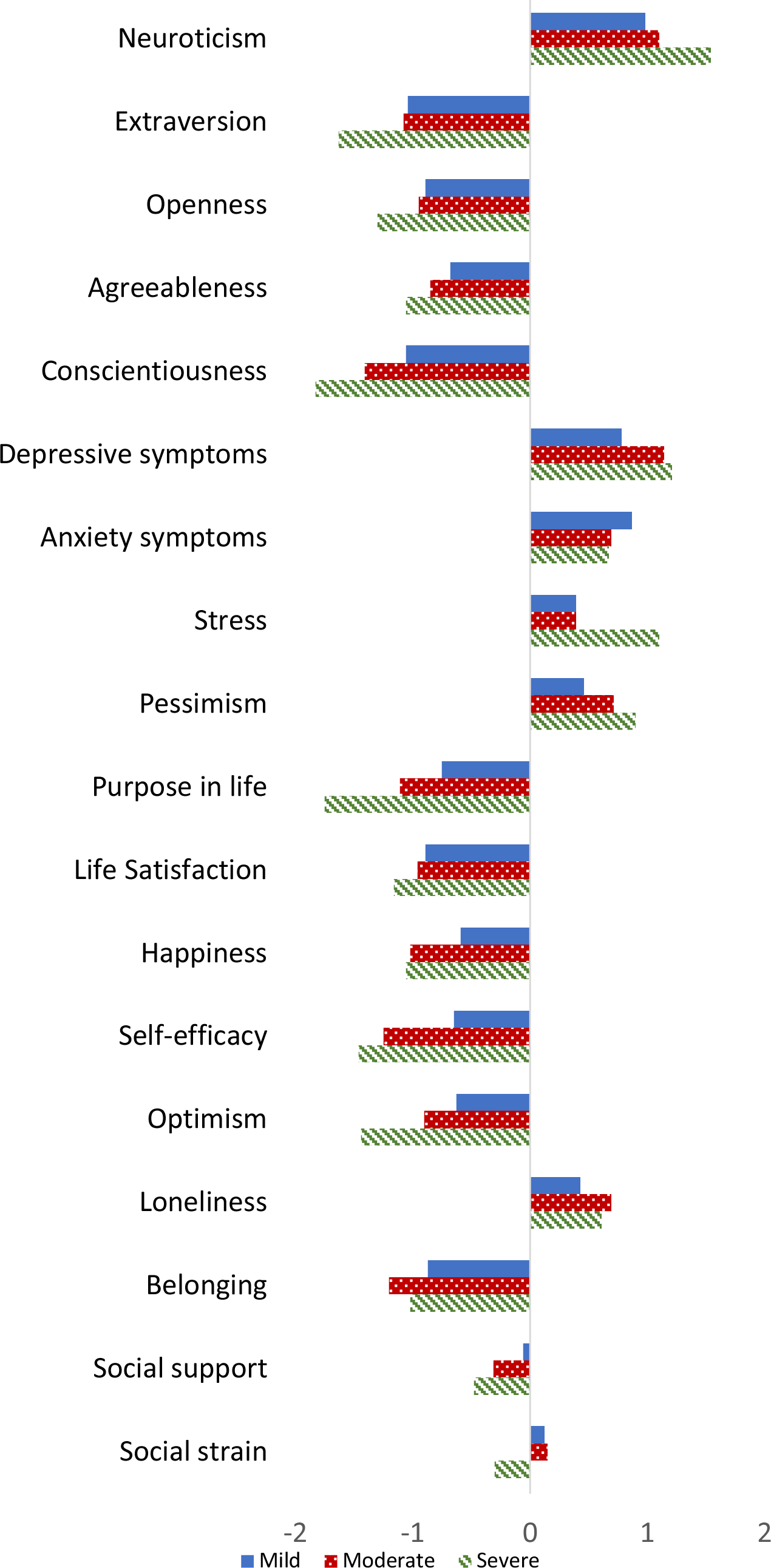
Change in personality (neuroticism, extraversion, openness, agreeableness, conscientiousness), psychological distress (depressive symptoms, anxiety symptoms, perceived stress, pessimism), psychological well-being (purpose in life, life satisfaction, happiness, self-efficacy, optimism), and social connection (loneliness, belonging, social support, social strain) by stage of dementia (mild, moderate, severe) quantified with Cohen’s d.

## Discussion

The present study applied a retrospective design that had been established previously to evaluate change in personality with dementia to examine whether a broad range of aspects of psychological function and social connection also changed with dementia. In a large sample of caregivers of people with dementia, we replicated the retrospective change in personality typically observed in people with dementia by their caregivers.^7–9^ Similar changes were observed across the measures of psychological function and social connection: Caregivers perceived that their care recipient increased in psychological distress and decreased in psychological well-being and social connection. Compared to mild dementia, the changes were larger in individuals with severe dementia on most outcomes.

The retrospective changes in FFM personality traits in the present study were consistent with a recent meta-analysis of personality change in dementia using the same approach.^11^ In particular, in the current sample, as in the meta-analysis, the largest changes were observed for neuroticism, extraversion, and conscientiousness (*d*>1.00). The increase in neuroticism and decrease in conscientiousness may reflect the growing inability to regulate the self emotionally and behaviorally, respectively. Areas of the brain responsible for effective self-regulation are among the first to deteriorate in dementia,^22^ and thus the clinical manifestation of that degeneration may be increases in sensitivity to negative emotions and decreases in the ability to plan and delay gratification. The decline in extraversion may likewise reflect a decline in the ability to regulate oneself socially. The social isolation that also often accompanies dementia^23^ may further hasten declines in extraversion.

The most retrospective changes observed in the current study among the measure of psychological distress were the increases in depression and anxiety. Depression and anxiety have long been implicated in dementia as risk factors,^24^ prodromal indicators,^25^ and symptoms after diagnosis.^26^ The present study suggests that caregivers perceive their care-recipient to be more prone to depression compared to how they were before their dementia developed. This difference is consistent with the higher prevalence of depression and the greater increases in depressive symptoms over time among individuals with dementia compared to older adults with normal cognition.^27^ Caregivers likewise perceived increases in other aspects of psychological distress and negative self-beliefs, particularly increases in perceptions of stress and a more pessimistic worldview.

Similar to psychological distress, but in the opposite direction, there were significant retrospective declines in all aspects of psychological well-being, particularly self-efficacy and purpose in life. Well-being is both a component of quality of life^28^ and a significant predictor of better cognitive outcomes.^29^ Purpose in life, for example, has gained attention recently because of its consistent association with better cognitive outcomes along the continuum of dementia:^30^ It is associated with healthier cognitive function prior to dementia,^31^ lower risk of pre-dementia syndromes,^32^ lower risk of incident dementia,^33^ and, even after the onset of dementia, fewer psychological and behavioral symptoms of dementia.^34^ Given the importance of purpose in life to well-being^35^ and that it can be maintained in dementia with more goal-directed activities,^36^ approaches to support purpose during dementia may help promote better quality of life.

There was also evidence that multiple aspects of social connection were disrupted with dementia. Specifically, caregivers perceived their care-recipient as less socially integrated (i.e., feeling lonelier and less belonging) with fewer supportive relationships and greater strain with others around them. Social connection is associated consistently with better cognitive outcomes.^37,38^ There is less evidence that specific dimensions of social health change with cognitive impairment. Self-reported loneliness, for example, tends not to change in the prodromal and early stages of dementia.^39^ It may be that declines in social connection may be more apparent later in the disease process and/or that individuals with dementia are not able to update their self-perceptions and thus may be more likely to report on how they would evaluate themselves prior to dementia. Caregivers may also report on aspects of social connection, particularly feelings of support and strain, through perceptions of their own feelings of support and strain for their care recipient.

Caregivers perceived changes in all aspects of psychological distress, well-being, and social connection included in this study. These changes could reflect, in part, the broad effects of neurodegeneration caused by dementia, which can disrupt regulation of emotion and behavior.^3^ Without effective regulation, individuals with dementia may be more prone to negative emotionality and difficulties with control over their behavior. Since this neurodegeneration is also responsible for the cognitive deficits critical for a diagnosis of dementia, it is not surprising that significant changes in psychological function and social connection would be apparent even in mild dementia. Aspects of behavioral control (e.g., conscientiousness) and future-oriented processes (e.g., purpose in life) may continue to be more vulnerable as the disease progresses.

The present study used an approach to identifying change that has been used in at least 18 studies of personality traits over the last 30 years.^11^ This approach, however, may exaggerate change with dementia. It is possible, for example, that caregivers may idolize their care recipient compared to their actual prior self (e.g., see the care recipient as more conscientious than they actually were) and, at the same time, they may likewise amplify the present (e.g., occasional specific behaviors that may be interpreted as low conscientiousness are perceived to mean that the individual is broadly low in conscientiousness). Such distorted perceptions may thus inflate the difference between past and current psychological function and social connection. These changes, however, are consistent with the clinical presentation of dementia^40^ as well as criteria for diagnosis.^2^ In addition, even if such changes are exaggerated, they are also the lived experience of the caregivers that may have implication for their care recipients. For example, caregivers may perceive their care recipients to be socially withdrawn and may not help them seek out social connection because they perceive that the care recipient is not interested in frequent social interactions anymore.

The present study had several strengths, including the relatively large sample of caregivers of individuals with dementia, the broad approach to psychological and social health, and the evaluation of change by stage of dementia. There are also limitations to consider. First, as described above, there may be biases in the method, including recall and contrast biases, that could exaggerate the differences observed from before to during dementia. Second, the type of dementia was not measured, so it was not possible to examine differences across dementia type (e.g., Alzheimer’s disease, vascular dementia, frontotemporal dementia, etc.). Third, the sample was nearly entirely White, and all participants were living in the United States. Future research would benefit from more diverse samples, including samples from other areas of the world, to be able to evaluate generalizability of the observed changes. Despite these limitations, the present study provides novel information on how aspects of psychological distress, well-being, and social connection change with dementia, in addition to personality traits, and quantifies the changes that are often observed clinically on the natural history of the disease.

## Data Availability

Data for analyses reported in this manuscript are available publicly at https://osf.io/3mg82/?view_only=0193b4e5c89245c3b49b0a0f0822a90b

## Acknowledgements

Research reported in this publication was supported by the National Institute on Aging of the National Institutes of Health under Award Numbers R01AG053297, R01AG074573, and R01AG068093. The content is solely the responsibility of the authors and does not necessarily represent the official views of the National Institutes of Health. The authors have no conflicts of interest to report. Data for analyses reported in this manuscript are available publicly at https://osf.io/3mg82/?view_only=0193b4e5c89245c3b49b0a0f0822a90b.

---

**Supplemental Table S1.**
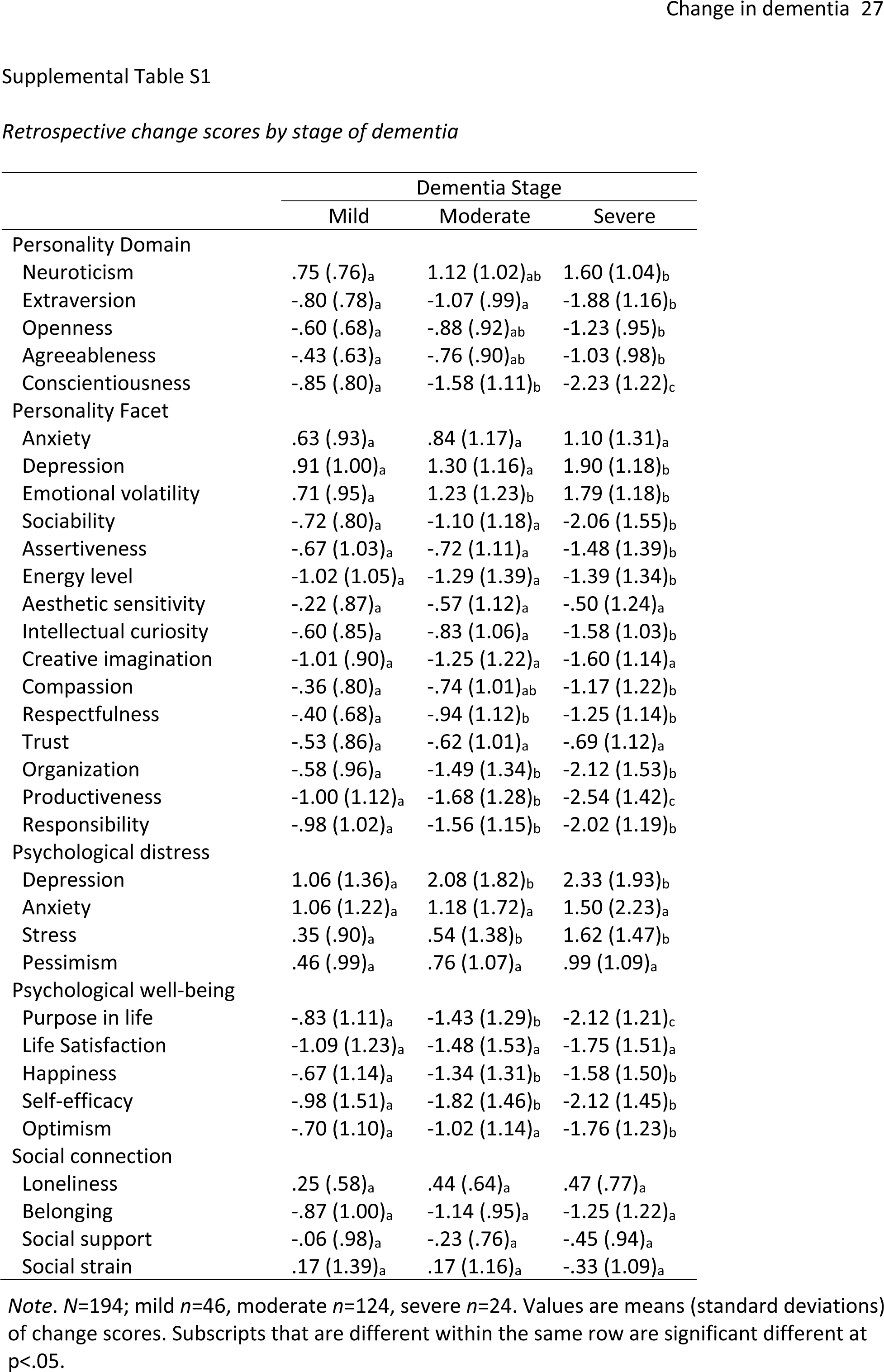
Retrospective change scores by stage of dementia.

## Example Survey

Example survey for ratings of personality, psychological distress, well-being, and social connection. Caregivers rated the person with dementia twice: once as they were prior to dementia and once as they are currently. The order was randomized across participants. The example survey is worded with the “he” pronoun. There were also “she” and “they” versions; caregivers chose the preferred pronoun for their care recipient.

Q27 We would like to know about the person you care for as he is **<u>NOW</u>**and how he was **<u>BEFORE</u>** he had dementia. You will be asked to answer the same questions twice. Please do not skip any questions, even if the questions seem repetitive.

Q28 **Please rate the person you care for as he was <u>BEFORE</u> he developed dementia.**

Q29 Here are a number of characteristics that may or may not apply to him **<u>before</u>** he developed dementia. For example, was he someone who liked to spend time with others? Please rate the extent to which you agree or disagree with each statement for how he was **<u>prior</u>** to the development of dementia:

He was someone who…

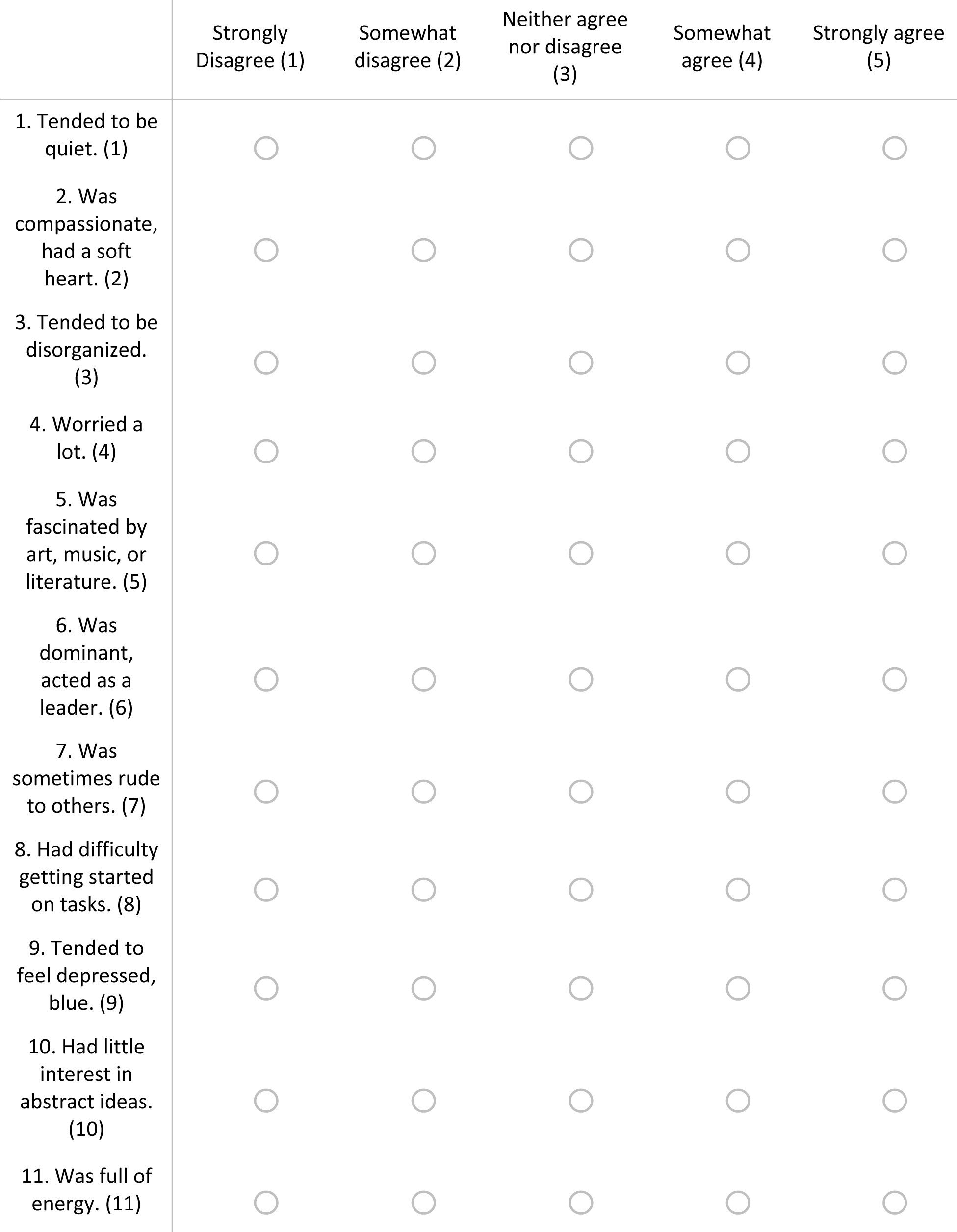
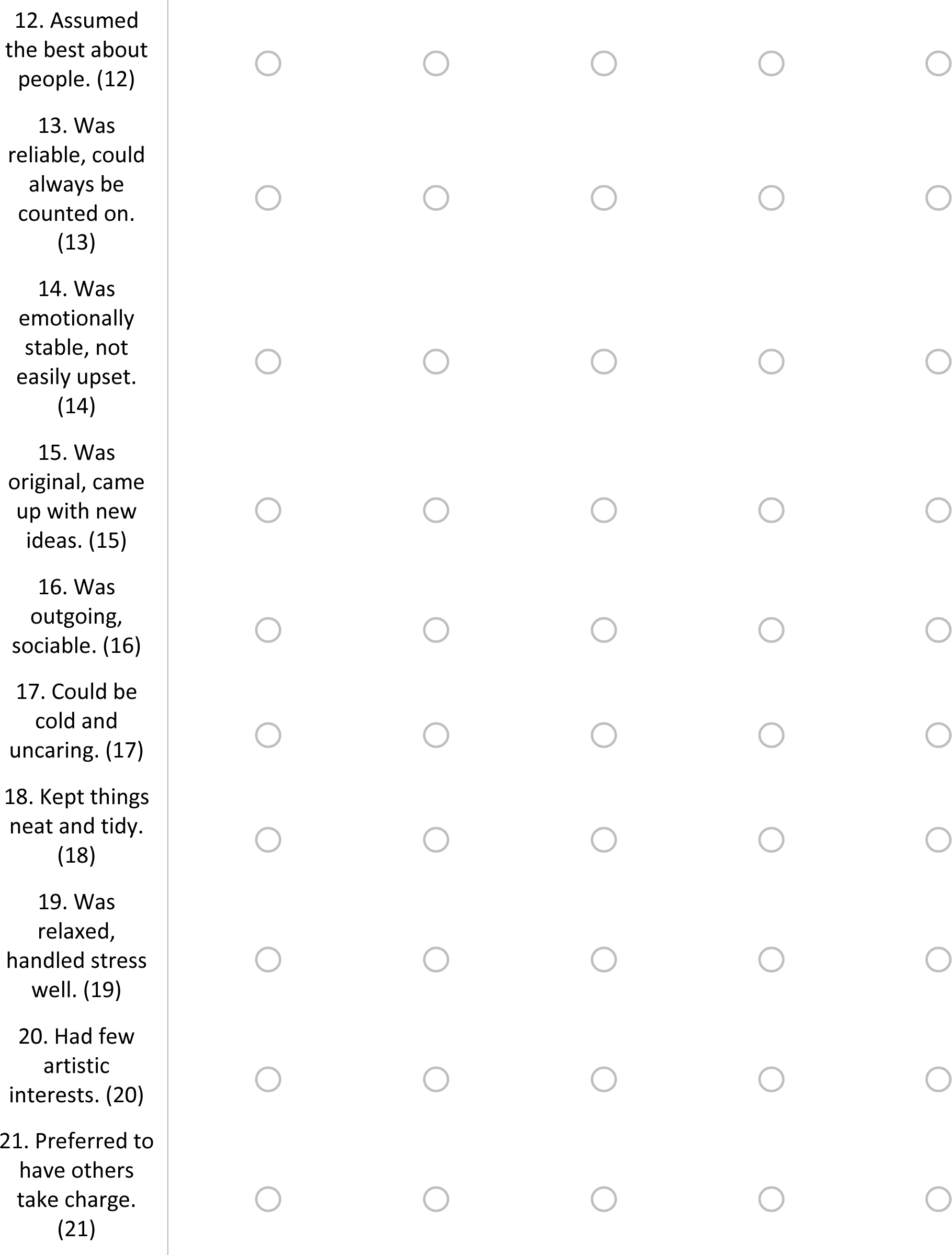
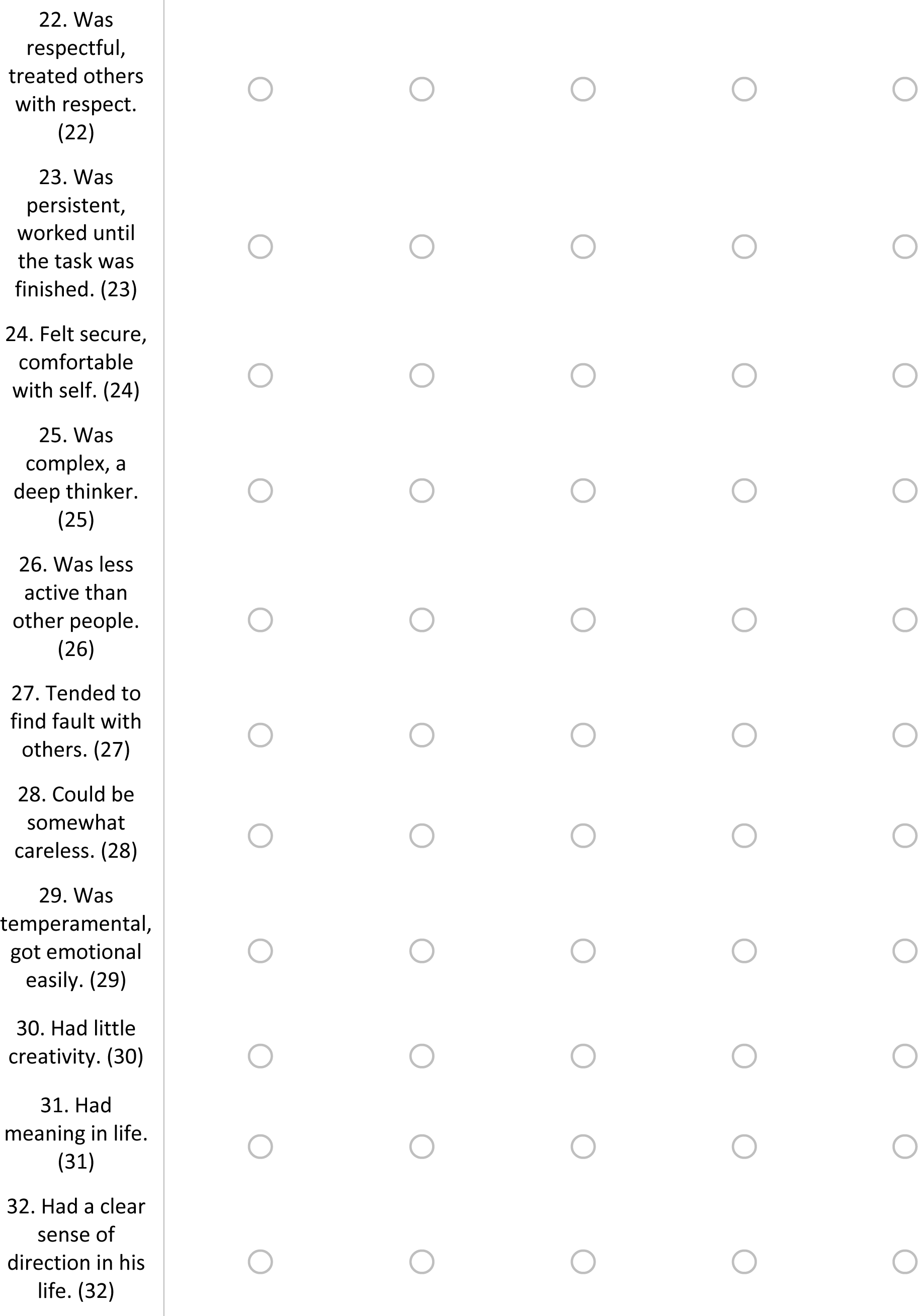
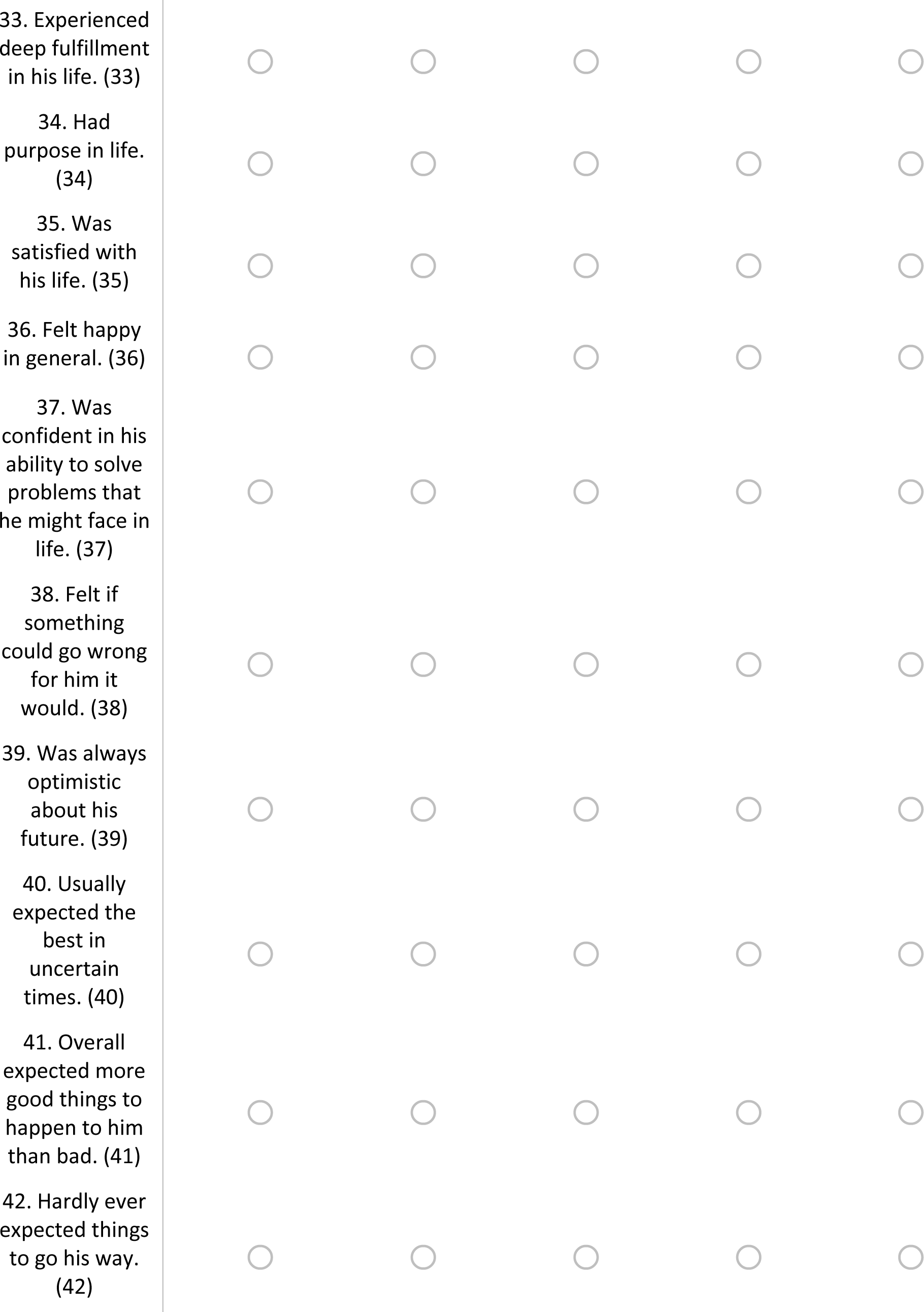
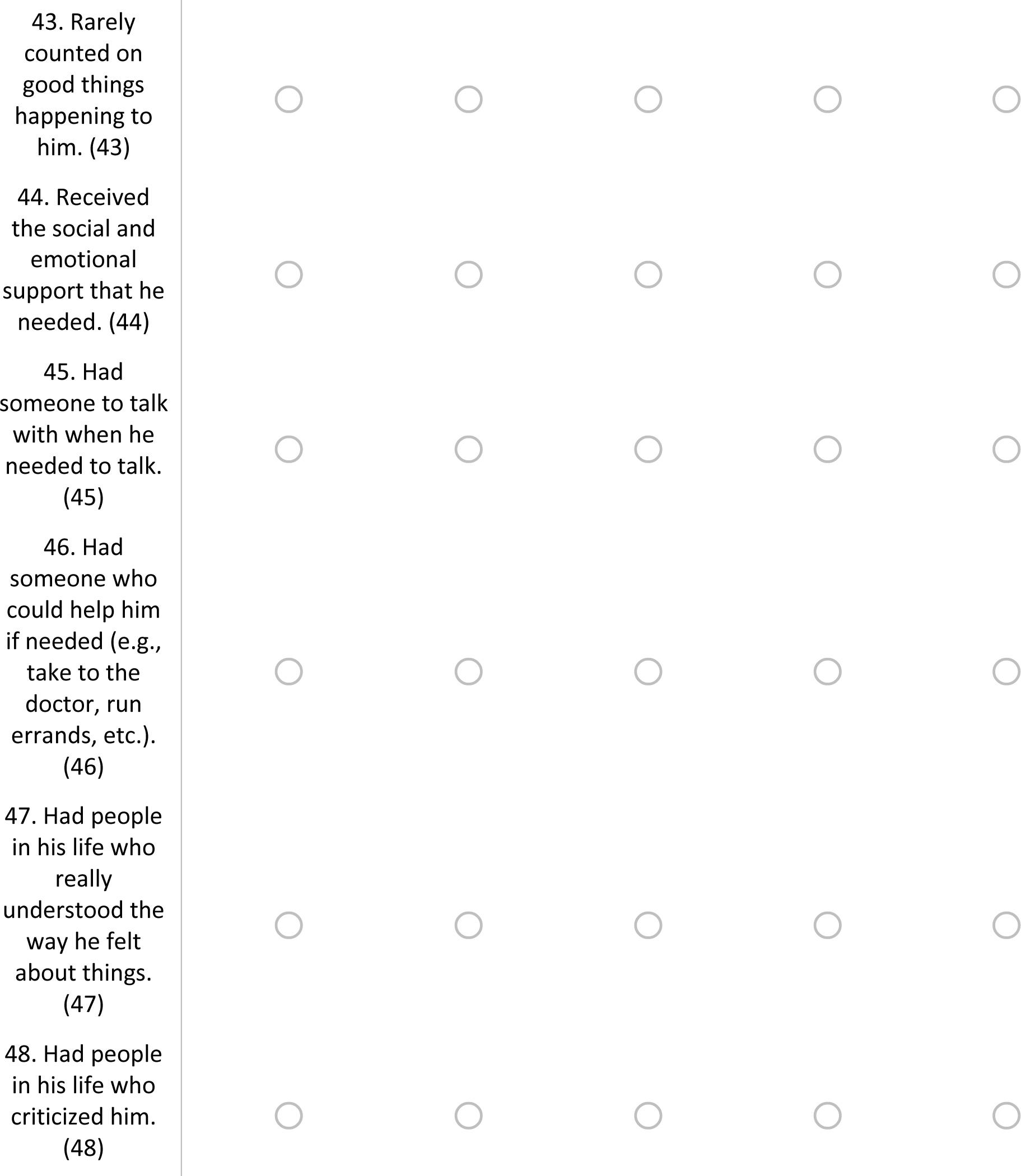

Q30 The next questions are about how he felt about different aspects of his life **before** dementia. How much of the time did he feel…

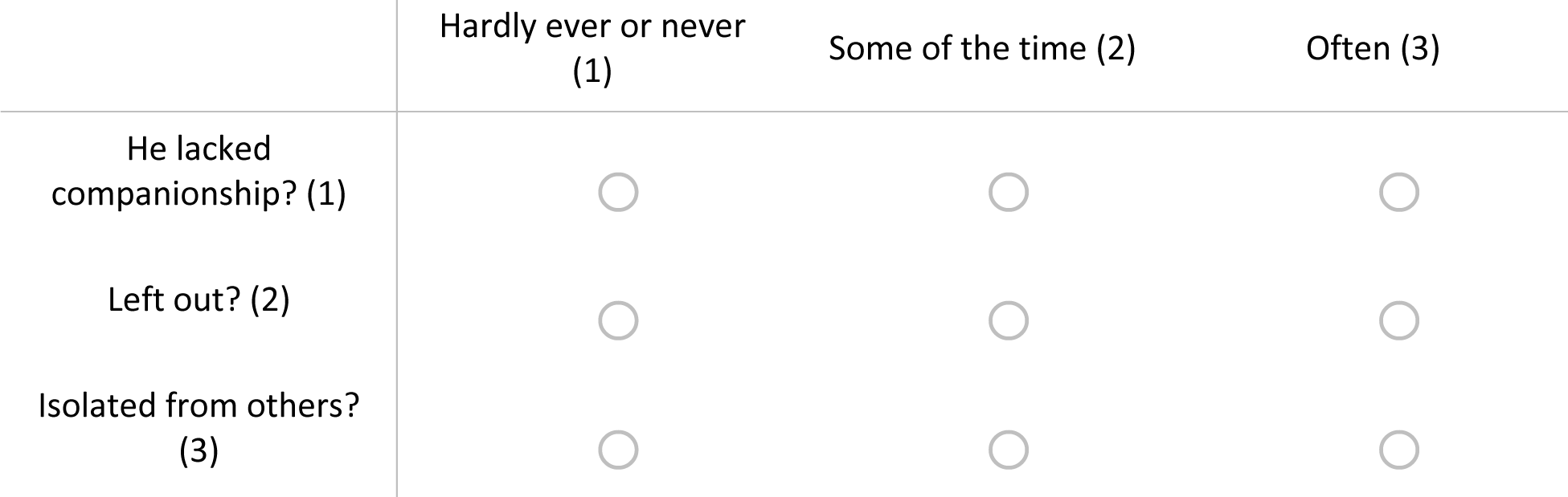

Q31 How often was he bothered by the following problems **before** dementia?

|  | Not at all (0) | Some of the time<br>(1) | More than half<br>the time (2) | Nearly all the<br>time (3) |
| --- | --- | --- | --- | --- |
| Feeling nervous, anxious or on edge. (1) | ☐ | ☐ | ☐ | ☐ |
| Not being able to stop or control worrying. (2) | ☐ | ☐ | ☐ | ☐ |
| Feeling down, depressed or hopeless. (3) | ☐ | ☐ | ☐ | ☐ |
| Little interest or pleasure in doing things. (4) | ☐ | ☐ | ☐ | ☐ |

Q32 How would he rate the amount of stress in his life **<u>before</u>** dementia?

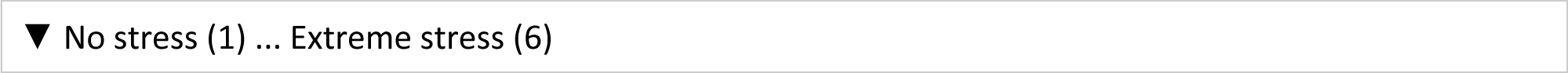

Q33 How would he describe his sense of belonging to his local community **<u>before</u>** dementia? Would he say it was:

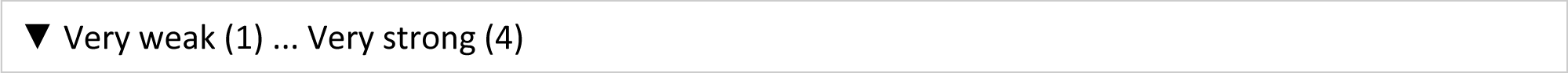

Q34 **This section refers to how he is <u>NOW</u>.**

Q35 Here are a number of characteristics that may or may not apply to him **<u>now</u>**. For example, is he someone who likes to spend time with others? Please rate the extent to which you agree or disagree with each statement for how he is **<u>now</u>**:

He is someone who…

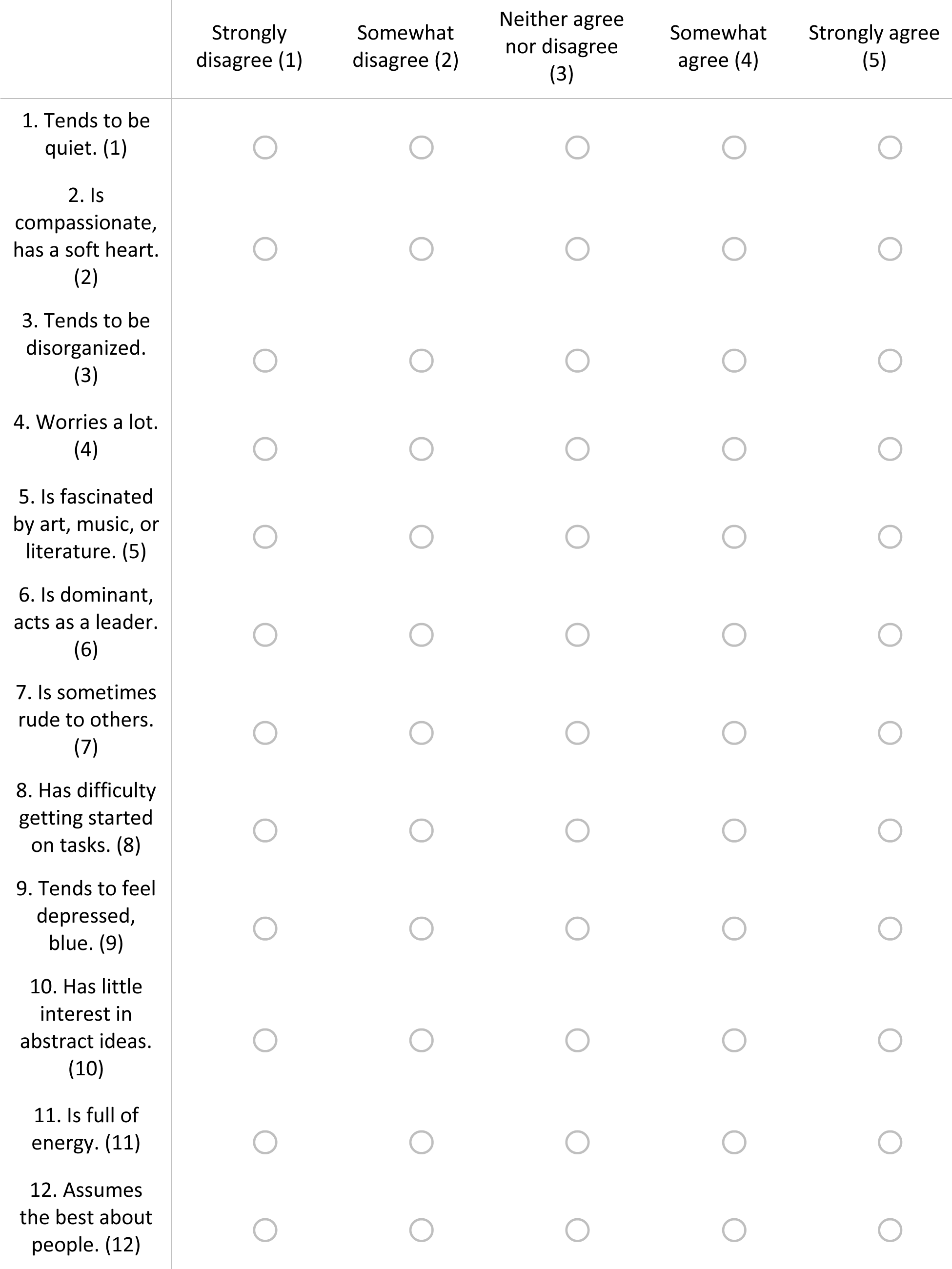
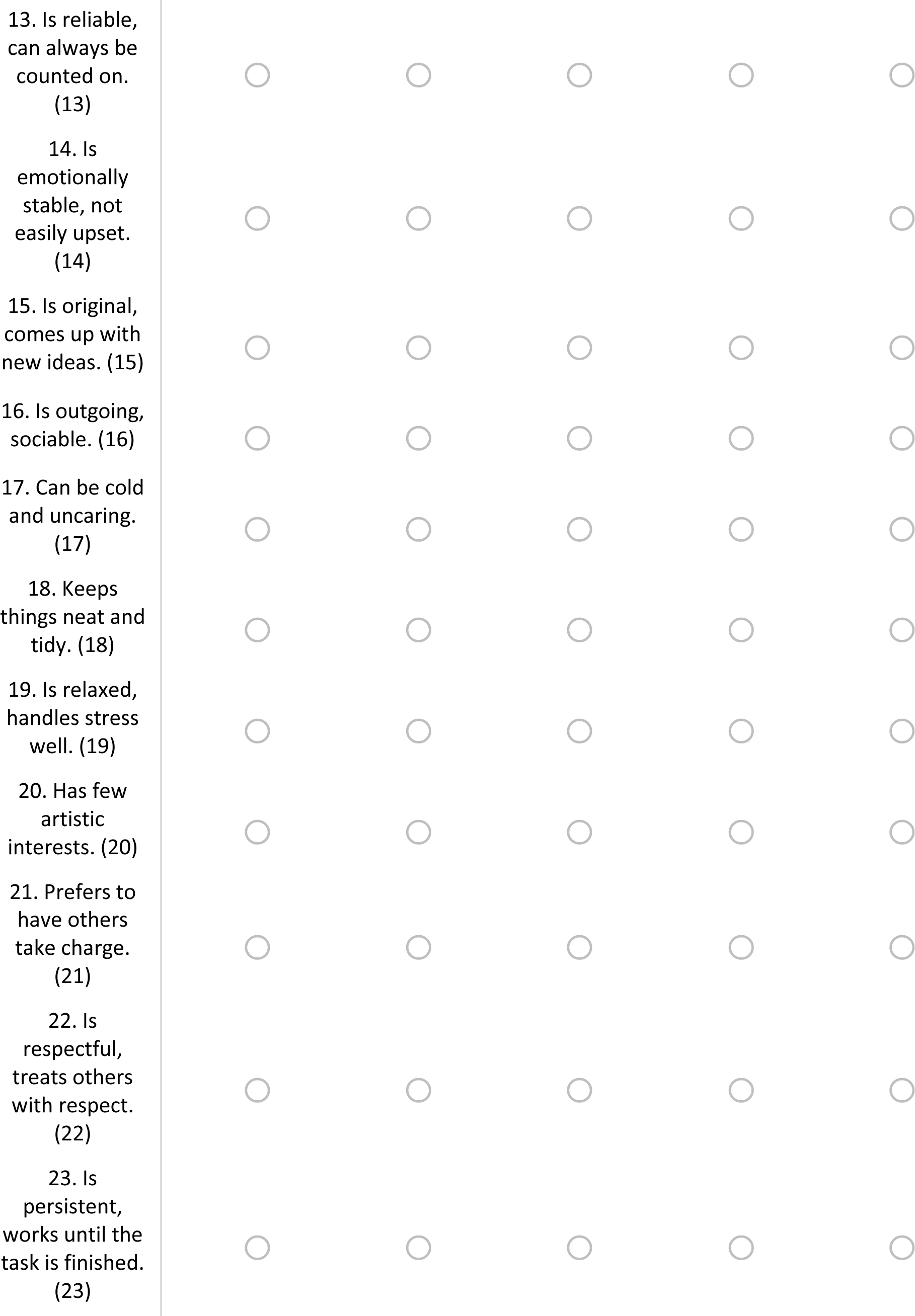
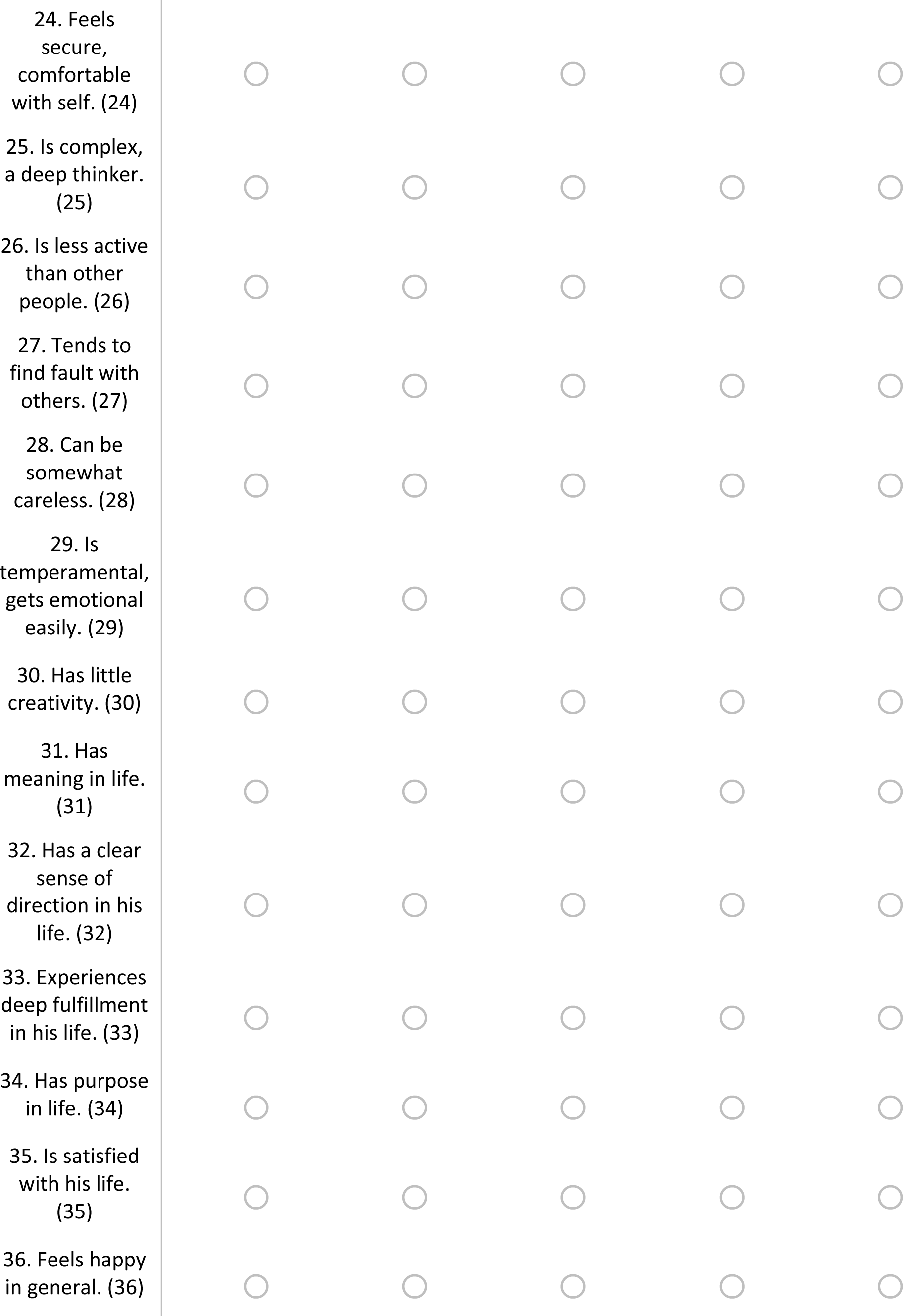
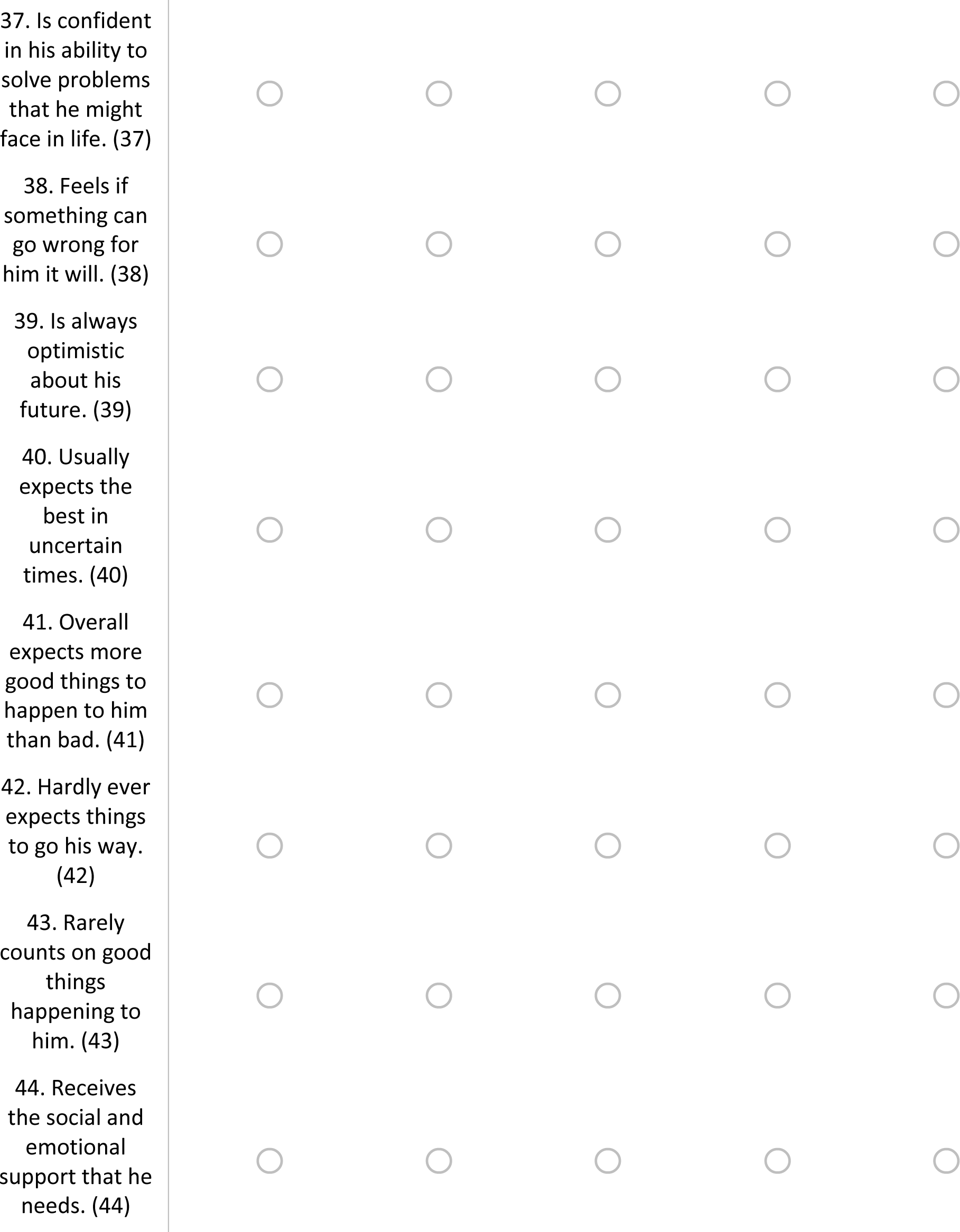
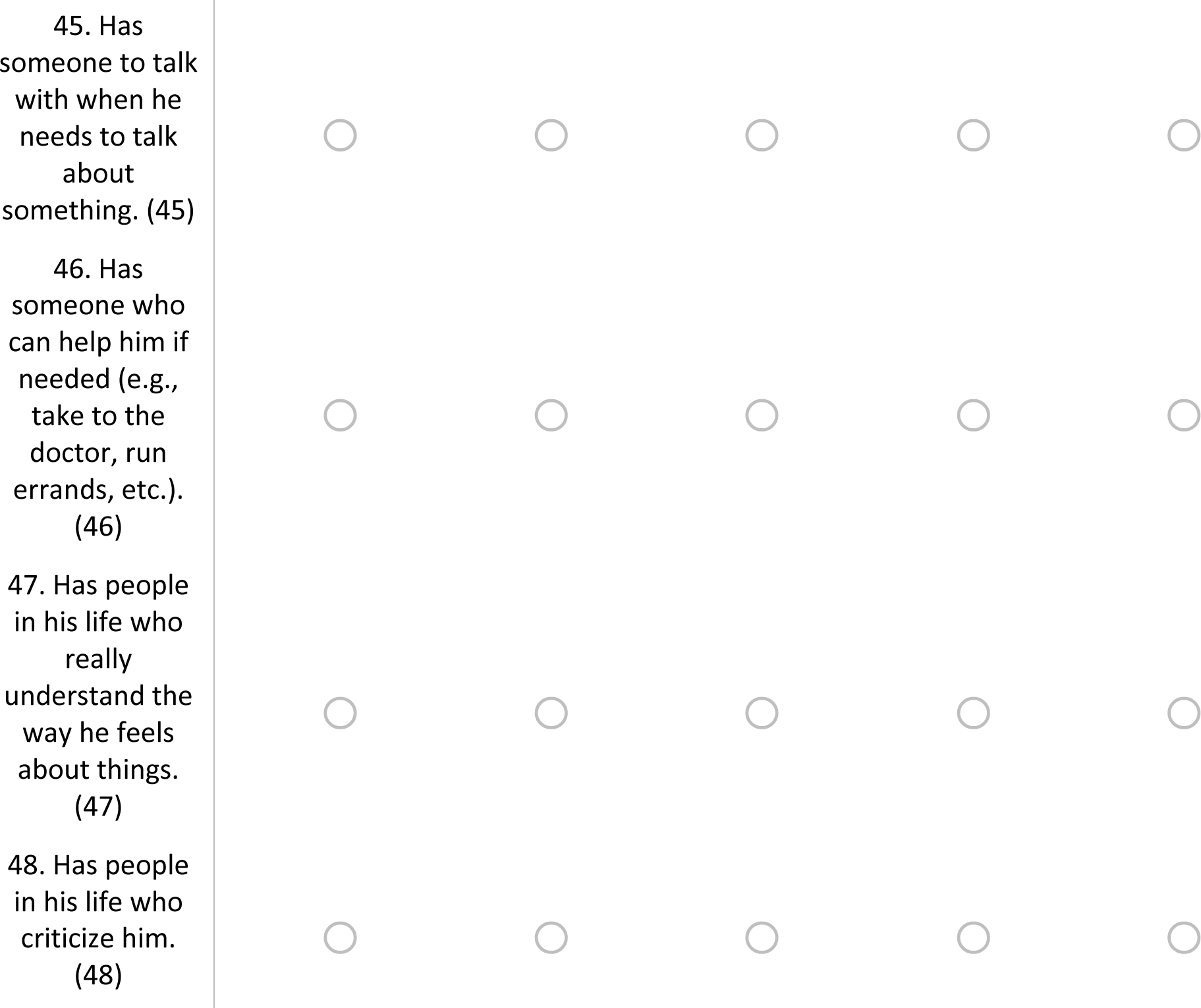

Q36 The next questions are about how he feels **<u>now</u>** about different aspects of his life. How much of the time does he feel…

|  | Hardly ever or never<br>(1) | Some of the time (2) | Often (3) |
| --- | --- | --- | --- |
| He lacks companionship? (1) | ☐ | ☐ | ☐ |
| Left out? (2) | ☐ | ☐ | ☐ |
| Isolated from others? (3) | ☐ | ☐ | ☐ |

Q37 How often has he been bothered **<u>now</u>** by the following problems? Feeling nervous,

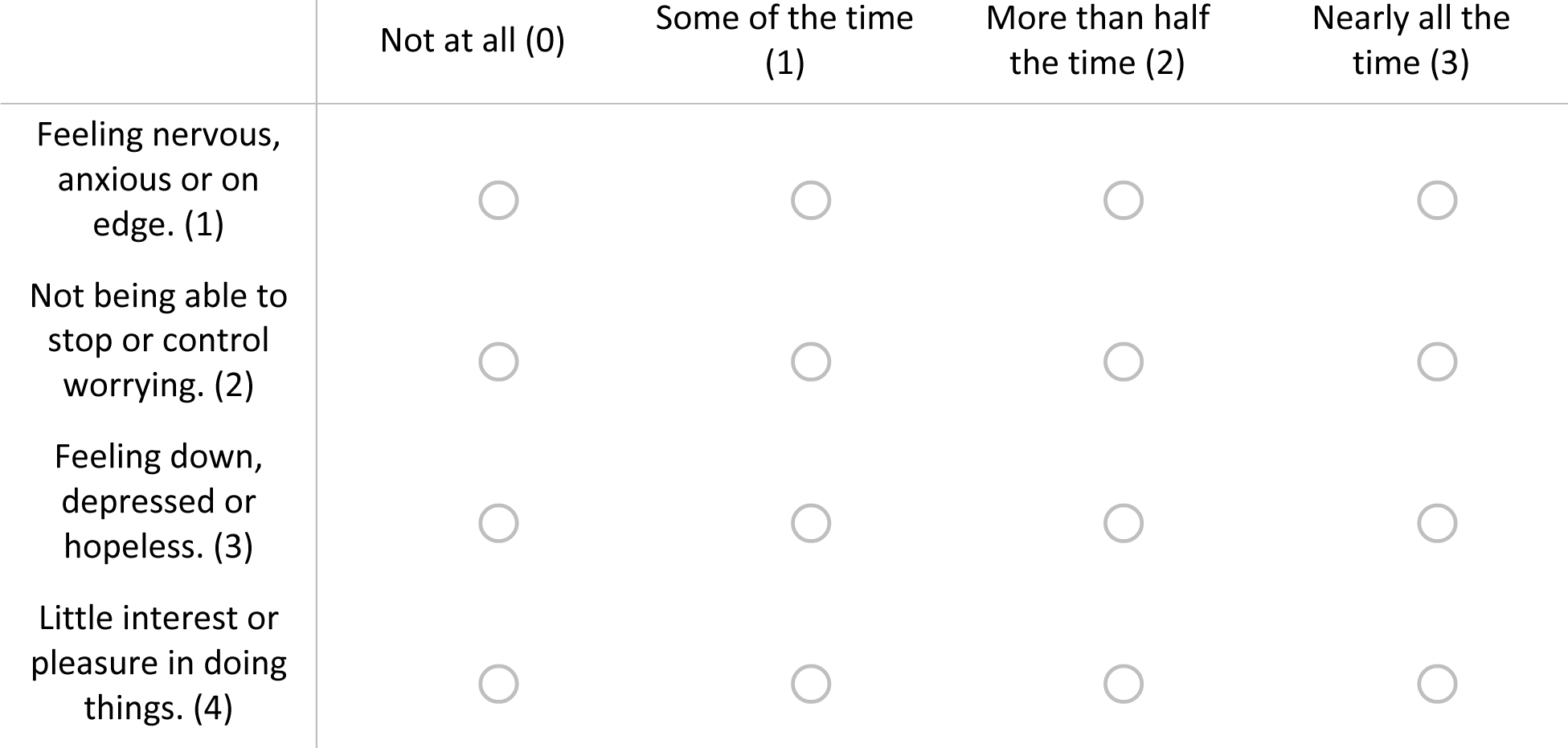

Q37 How often has he been bothered **now** by the following problems?
|  | Not at all (0) | Some of the time<br>(1) | More than half<br>the time (2) | Nearly all the<br>time (3) |
| --- | --- | --- | --- | --- |
| Feeling nervous,<br>anxious or on<br>edge. (1) | ☐ | ☐ | ☐ | ☐ |
| Not being able to<br>stop or control<br>worrying. (2) | ☐ | ☐ | ☐ | ☐ |
| Feeling down,<br>depressed or<br>hopeless. (3) | ☐ | ☐ | ☐ | ☐ |
| Little interest or<br>pleasure in doing<br>things. (4) | ☐ | ☐ | ☐ | ☐ |

Q38 How would he rate the amount of stress in his life **<u>now</u>**?

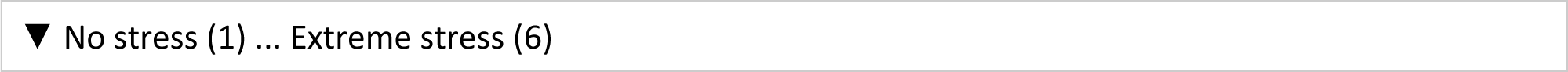

Q39 How would he describe his sense of belonging to his local community **<u>now</u>**? Would he say it is:

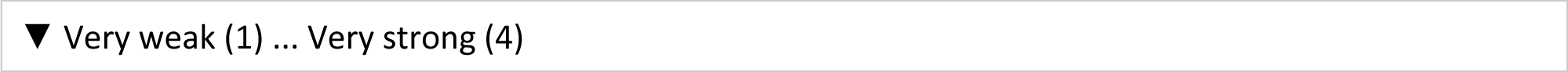

